# Calibrated per-pixel uncertainty for low-dose paediatric chest-radiograph denoising at no fidelity cost

**DOI:** 10.64898/2026.09.14.26363026

**Authors:** Samuel Tekle Mengistu, Kyriakos Flouris

**Affiliations:** Department of Computer Science, University of Oxford, Oxford, United Kingdom; Oxford Martin School, University of Oxford, Oxford, United Kingdom; The Accelerate Hub, University of Oxford, Oxford, United Kingdom; MRC Biostatistics Unit, University of Cambridge, Cambridge, United Kingdom

**Keywords:** image denoising, uncertainty quantification, calibration, variational autoencoder, paediatric chest radiography, low-dose imaging

## Abstract

**Background:** Deep denoisers restore low-dose chest radiographs but emit a single point estimate with no indication of where the output is reliable, so a confident-looking reconstruction can be locally wrong. In paediatric radiography, where the radiation-dose imperative is sharpest, we ask whether a per-pixel uncertainty map can be attached at no meaningful cost to reconstruction quality.

**Methods:** We develop PP-VAE-Hformer, a hybrid convolution–transformer denoiser with a variational-autoencoder bottleneck (epistemic uncertainty map) and a heteroscedastic dual head (aleatoric map), and run a 19-arm ablation isolating five composite-loss terms on Poisson–Gaussian-degraded paediatric chest radiographs (Kermany collection; 624-image held-out test set), against eight discriminative baselines retrained on identical data and noise. Calibration is assessed by reliability diagrams and post-hoc *σ*-scaling; comparisons use Bonferroni-corrected Welch tests with Cohen’s *d* as the primary discriminant.

**Results:** A heteroscedastic negative-log-likelihood (NLL) objective buys a calibrated aleatoric map for 1.2 dB PSNR, which two-stage fine-tuning recovers to a statistically indistinguishable 0.010 dB. For the variational variant the Kullback–Leibler (KL) annealing *schedule*, not the bottleneck itself, is decisive: cyclic annealing closes a 1.10 dB gap over posterior-collapsing linear warmup. Predicted uncertainty tracks realised error (Pearson *r >* 0.8) with an optimal recalibration scale within 1% of unity. A matched-severity cross-degradation test shows this calibration is model-circular, however: at equal severity the aleatoric map becomes modestly (8–16%, a lower bound) over-confident once the noise *model* changes, and the Monte-Carlo epistemic map is a weaker error predictor that does not compensate. We additionally document a previously unreported destructive interaction between structural-similarity and Sobel-edge supervision at literature-default weights.

**Conclusions:** A calibrated per-pixel uncertainty map can be attached to a paediatric low-dose CXR denoiser at effectively no reconstruction cost. Its near-perfect calibration is tied to the trained (“inverse-crime”) noise model, and the Monte-Carlo epistemic map does not reliably substitute where the aleatoric map fails, so an architecture-independent uncertainty estimate (e.g. deep ensembling), a blinded reader study, and real low-dose data are the immediate next steps.

## 1 Background

Chest radiography is the most common paediatric imaging examination, yet the ionising radiation that forms each image carries a measurable lifetime cancer risk that is steeper in children: large cohorts link paediatric imaging radiation to excess leukaemia and brain tumours under a no-safe-threshold model [1–3]. Lowering dose reduces this risk but raises signal-dependent quantum noise, and by the Rose criterion a contrast-to-noise ratio of roughly five is needed before a human observer can reliably detect a low-contrast finding [4, 5]; dose reduction erases faint parenchymal detail long before it touches the high-contrast bony thorax. Every noisy image that forces a repeat acquisition, or an escalation to chest CT at a far greater bone-marrow dose [3], turns that statistical risk into avoidable harm. Deep denoisers restore much of the lost quality, but from classical filters [6] through discriminative convolutional networks [7, 8] to hybrid transformers [9, 10] they output a single point estimate with no measure of where that estimate can be trusted, while generative denoisers [11, 12] can hallucinate anatomy [13, 14]. None provides a *calibrated* per-pixel uncertainty map that separates pixels recovered from the measurement from pixels filled in by a learned prior. In paediatric radiology (where the dose imperative is sharpest and documented generalisation failures of adult-trained models are a recognised safety concern [15]), this gap is concrete: a denoiser deployed to enable dose reduction should report, spatially, how far its own output can be trusted. Regulators already withhold trust from adult-derived evidence in this population: the WHO policy on computer-aided detection for tuberculosis screening restricts its recommendation to individuals aged 15 and older, explicitly excluding younger children for insufficient evidence [16], precisely the generalisation gap a spatially calibrated uncertainty map is positioned to flag.

We ask a single question: can replacing the pixel-wise loss with a heteroscedastic, structure- and frequency-aware objective, together with a variational bottleneck, equip a paediatric low-dose chest X-ray (CXR) denoiser with a per-pixel uncertainty map at no clinically meaningful cost to reconstruction quality? We answer it with a controlled 19-arm ablation of a fixed hybrid convolution–transformer backbone, evaluated against eight established discriminative denoisers retrained from scratch on identical data and noise, so that the ablation isolates the training *objective* and the baseline comparison isolates the *architecture*, both free of cross-paper confounds.

Our contributions are:

1. a single 19-arm ablation isolating all five composite-loss terms at once, where prior work tests them in isolation [17, 18], whose ranking holds under held-out perceptual metrics (FSIM, LPIPS) that no arm trains on;
2. the finding that the KL-annealing *schedule*, not the variational bottleneck itself, is the decisive design variable for VAE-based denoising (1.10 dB, cyclic vs. linear warmup), and that two-stage loss-substitution fine-tuning erases the 1.2 dB calibration tax an uncertainty head otherwise costs, to a statistically indistinguishable 0.010 dB;
3. a previously unreported destructive interaction between structural similarity (SSIM) and Sobel-edge supervision at literature-default weights (−1.75 dB, unrecovered by any re-weighting), invisible to either term alone, together with a second guardrail result that PReLU activations destabilise heteroscedastic variance heads; and
4. a calibration analysis showing the aleatoric map is well calibrated at the pixel level (*σ*-scaling within 1% of unity), together with a matched-severity cross-degradation test that makes the inverse-crime caveat [19] concrete: the calibration is tied to the trained noise model, and the Monte-Carlo epistemic map does not reliably compensate where the aleatoric map fails.

### 1.1 Related work

#### Denoising architectures

Learned denoising progressed from residual CNNs [7, 20, 21] to deep noise-conditioned U-Nets [8], windowed-attention transformers [9, 22], gated convolutional designs [23], and Swin–convolution hybrids trained on realistic noise [24]; Hformer [10] interleaves convolution and attention for low-dose CT. For paediatric CXR specifically, SharpXR [25] pairs a denoising decoder with a Laplacian edge branch. All are deterministic: none attaches a reliability signal to its output.

#### Loss functions

Pixel norms (L2, L1, Charbonnier [26]) converge to the posterior mean and over-smooth [17]; structural [27, 28] and frequency-domain [29, 30] terms recover perceptual and spectral content that spectral bias suppresses [31]. Prior work evaluates these terms largely in isolation; interactions between them at literature-default weights are undocumented.

#### Uncertainty quantification

Heteroscedastic regression heads capture aleatoric (data) uncertainty and Monte-Carlo sampling of a stochastic representation captures epistemic (model) uncertainty [32, 33]; deep ensembles offer an architecture-independent alternative [34]. Variational autoencoders [35] supply a principled stochastic bottleneck but are prone to posterior collapse, mitigated by cyclical annealing [36, 37] or free bits [38]. Yeoh et al. [39] survey VAE applications in medical imaging; to the best of our knowledge, and consistent with that survey, no prior system combines hybrid-attention reconstruction with calibrated per-pixel aleatoric and Monte-Carlo epistemic maps on paediatric CXR.

## 2 Methods

### 2.1 Study design

Nineteen arms were defined on a single fixed backbone: sixteen core arms (A–P) each differing only in the loss function or one specific design choice (probabilistic vs. deterministic latent space, KL schedule, activation function), plus a Charbonnier pixel-norm variant (Q) trained from scratch and two fine-tuning arms (R, S) that resume from a converged checkpoint. Each arm adds exactly one component to its predecessor, so performance differences are attributable to the component under test [40]. Every model (the 19 arms and eight retrained baselines) is trained and evaluated on identical data and noise. Table 1 summarises the design; Fig. 1 shows the end-to-end pipeline.

**Figure 1:**
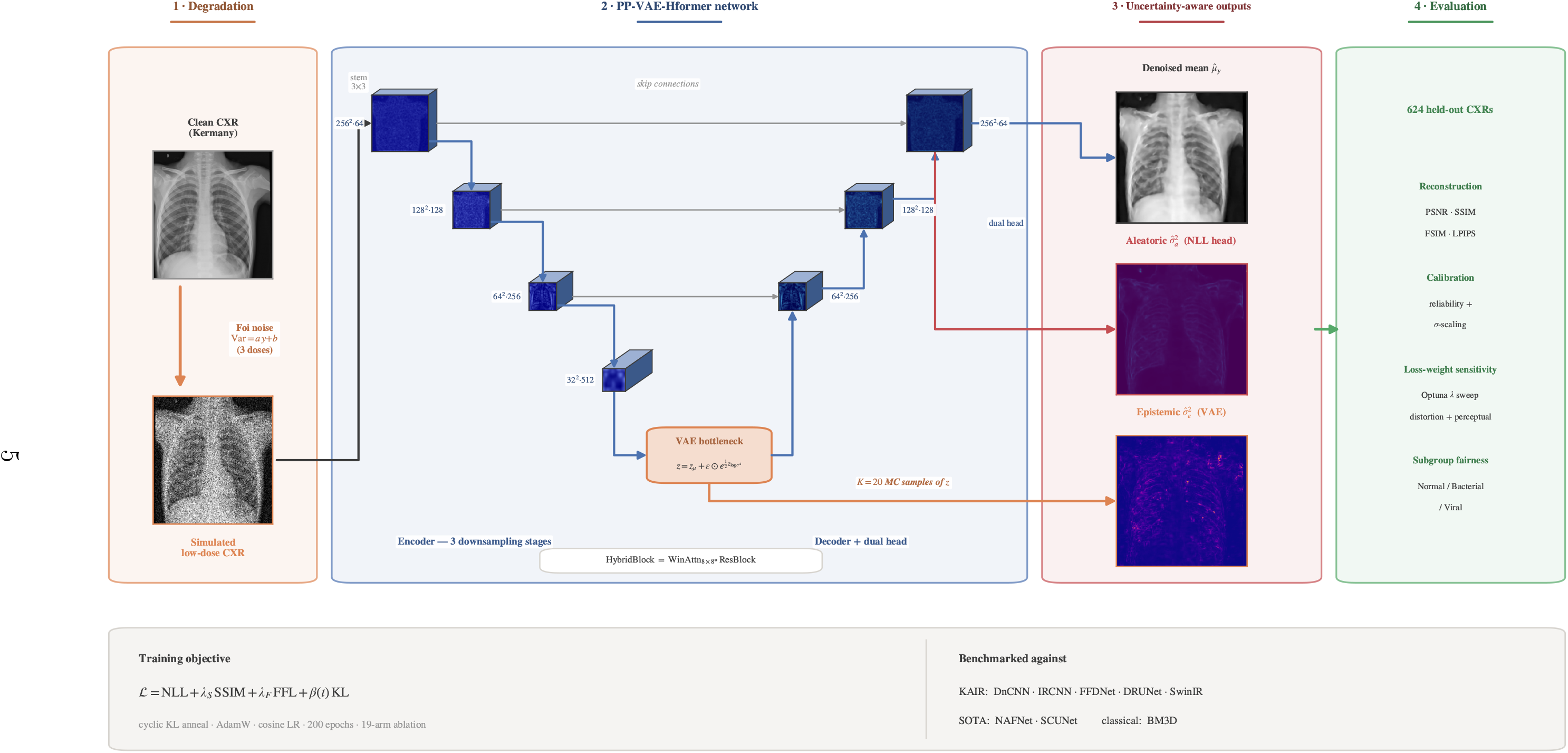
End-to-end study pipeline. (1) A clean Kermany CXR is degraded by the Foi noise model at three dose levels. (2) The noisy image passes through the PP-VAE-Hformer U-Net with an optional VAE bottleneck. (3) The dual head outputs the denoised mean *µ̂*_y_ and aleatoric variance 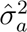; the epistemic variance 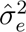 comes from *K*=20 Monte-Carlo samples. (4) Evaluation is on 624 held-out CXRs against eight retrained baselines.

**Table 1:** Active loss terms and design features for all 19 ablation arms. All other settings (architecture, optimiser, learning rate, data, schedule) are identical across arms.

| Arm | Objective / design variable | VAE |
| --- | --- | --- |
| A | MSE baseline (replicates DnCNN/Hformer training) | – |
| B | Heteroscedastic NLL | – |
| C | NLL + MS-SSIM | – |
| D | NLL + MS-SSIM + FFL | – |
| E | NLL + SSIM + FFL + KL, linear warmup | ✓ |
| F | as E, cyclical KL annealing | ✓ |
| G | as E, free-bits KL | ✓ |
| H | as E, cyclical + free-bits KL | ✓ |
| I | L1 | – |
| J | L1 + SSIM + FFL | – |
| K | NLL + L1 | – |
| L | NLL + Sobel edge + FFL | – |
| M | NLL + SSIM + Sobel edge + FFL | – |
| N | NLL + VGG-16 perceptual + SSIM + FFL | – |
| O | as D, PReLU decoder activations | – |
| P | as H + edge, PReLU activations | ✓ |
| Q | Charbonnier pixel norm, from scratch | – |
| R | two-stage FT from A: L1+SSIM+FFL | – |
| S | two-stage FT from A: NLL+SSIM+FFL | – |

### 2.2 Poisson–Gaussian degradation

We simulate low dose with the Foi affine-variance model [41], Var(*ỹ* | *y*) = *a y* + *b*, applied on-the-fly to displayed intensity *y* ∈ [0, 1] using a Gaussian approximation to the Poisson term, *ỹ* = *y* + *ε*, *ε* ∼ *N* (0, *a* max(*y,* 0) + *b*), clipped to [0, 1]. Three presets probe mild, moderate and severe noise ((*a, b*) = (0.01, 0.002), (0.03, 0.005), (0.08, 0.010); effective *σ* ≈ 15, 25, 50 at mean intensity, matching the standard evaluation grid [7]). Training noise is freshly sampled each epoch with the preset drawn uniformly per batch; validation and test noise is fixed per deterministic seed. The implementation was validated against its specification: 4 × 10^4^ realisations at each of 51 intensity levels recover the specified (*a, b*) to within 1% with *R*^2^ *>* 0.999 at every preset.

The affine variance law holds for processed X-ray images, with its parameters recoverable from transform-domain statistics [42] and the image-domain form adopted by recent paediatric denoisers [25]; here, because displayed brightness is inverted relative to photon flux, places noise on the bright bony thorax rather than the photon-starved lung. The noise remains signal-dependent and identical for every arm, so within-study comparisons are unaffected; the choice is this study’s principal degradation limitation (Section 4.1).

### 2.3 Architecture

PP-VAE-Hformer keeps the reconstruction backbone of Hformer [10] (a hybrid U-Net whose stages interleave a residual convolution branch with self-attention), realising the attention branch as non-overlapping 8 × 8 windowed self-attention following Swin [22], and adds the two components that make the output probabilistic. A 3 × 3 stem maps the 1×256×256 input to 64 channels; three encoder stages of two HybridBlocks each (ResBlock with GroupNorm [43] and GELU [44], residual skip [45], followed by windowed multi-head attention [46], *h* = 4) halve resolution and double channels to a 512×32×32 bottleneck; the decoder mirrors this with transposed convolutions and fused skip connections. Windowed attention keeps cost linear in pixels and the network compact at 25.7 M parameters (24.9 M without the VAE bottleneck), well below several baselines. All weights are trained from scratch; no pretraining or transfer learning is used.

#### VAE bottleneck and epistemic map

For the variational arms, three 1×1 convolutions at the bottleneck produce the posterior mean **z**_µ_, log-variance **z**_log σ^2^_, and the projection of the reparameterised sample 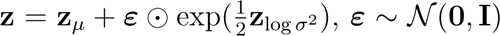 [35]. At evaluation, reconstruction uses the deterministic pass **z** = **z**_µ_; the epistemic map is the pixel-wise variance of *K*=20 stochastic passes, which under the law of total variance complements the aleatoric term [33].

#### Dual output head

Two parallel 3 × 3 heads at full resolution emit the reconstruction mean *µ̂*_y_ and the per-pixel log aleatoric variance log 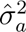 in a single forward pass.

### 2.4 Composite objective

The full objective is

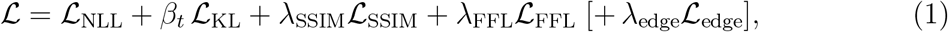

with heteroscedastic NLL [32]

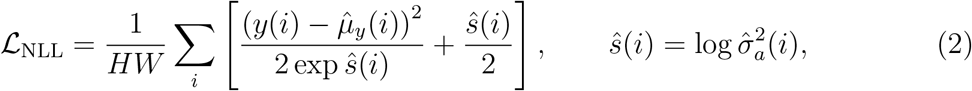

multi-scale SSIM [28] (weight *λ*_SSIM_ = 0.5; mixing a pixel and a structural term follows 17), focal frequency loss [29] (*λ*_FFL_ = 0.1, focal exponent *α* = 1), an optional gradient-difference edge term in the spirit of Mathieu et al. [47], here computed with Sobel filters (*λ*_edge_ = 0.1; Arms L, M, N, P), and the Gaussian KL divergence [48] in the per-dimension closed form of Kingma and Welling [35, App. B]. Arm E anneals *β* linearly to *β*_max_ = 10*^−^*^3^ over 20 epochs; Arm F uses cyclical cosine annealing over four 50-epoch cycles [36]; Arm G clips per-channel KL below *λ* = 0.25 nats (free bits [38]); Arm H combines both.

#### Two-stage loss-substitution fine-tuning

Arms R and S load Arm A’s converged L2 checkpoint and substitute a richer composite loss for 100 further epochs (R: L1+SSIM+FFL, *η* = 5 × 10*^−^*^5^; S: NLL+SSIM+FFL, *η* = 2 × 10*^−^*^5^, with a 20-epoch linear loss blend to stabilise the variance head), testing whether the NLL fidelity cost is recoverable without retraining.

### 2.5 Data

The Kermany paediatric CXR collection [49] comprises 5,856 anterior–posterior radiographs from children aged one to five years. Images were converted to greyscale, resized to 256×256 with antialiased bilinear interpolation, and normalised to [0, 1]. The 624-image test partition (234 Normal, 242 Bacterial, 148 Viral Pneumonia) was locked before development; of the remainder, 785 images (15%) formed the validation set (checkpoint selection only) and 4,447 the training set. Spatial augmentation (horizontal flip, rotation ±5°) was applied to training only; intensity augmentation was excluded as it would invalidate the calibrated noise parameters. Class labels were unused during training.

### 2.6 Baselines

Eight established discriminative denoisers were retrained from scratch on the identical split and noise model: DnCNN [7], IRCNN [20], FFDNet [21], DRUNet [8], SwinIR [9], NAFNet [23], SCUNet [24], and the paediatric-CXR-specific SharpXR [25] (under its authors’ RMSE objective); classical BM3D [6] with oracle noise knowledge completes the set. The first seven share the ablation’s training protocol. Because every baseline is retrained within-study, absolute figures are directly comparable to each other rather than to the original papers.

### 2.7 Training

All arms trained for 200 epochs, batch size 16, AdamW [50] at 2 × 10*^−^*^4^ with cosine annealing to 10*^−^*^6^, weight decay 10*^−^*^4^, mixed precision [51], and gradient clipping at 1.0, on NVIDIA A100 80 GB GPUs (Cambridge CSD3; ≈9–10 h per proposed arm, ≈398 GPU-hours total). The best validation-PSNR checkpoint was selected per arm.

### 2.8 Evaluation and statistical analysis

Reconstruction is scored by PSNR, SSIM and MS-SSIM; the held-out FSIM [52] (primary) and LPIPS [53] appear in no training objective and serve as unbiased perceptual judges. Calibration uses reliability diagrams (test pixels binned by predicted *σ̂*_a_ against realised RMSE, summarised by Pearson *r*), NLL, sharpness, and a post-hoc *σ*-scaling factor *s*^⋆^ fitted on held-out pixels [54]. Values are means over the 624-image test set with 95% bootstrap confidence intervals (1,000 resamples) [55].

Because the central claims are claims of *equivalence*, the analysis is built around effect magnitude. An omnibus ANOVA gates 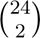 = 276 pairwise Welch *t*-tests [56] (robust to the strong heteroscedasticity between stable and failed arms), Bonferroni-corrected to *α*_adj_ = 1.81 × 10*^−^*^4^ [57]. At *n* = 624 nearly every pair reaches nominal significance [58], so Cohen’s *d* [59] is the primary discriminant (|*d*| *<* 0.2 negligible). Subgroup fairness across diagnostic classes uses Welch ANOVA with Bonferroni-adjusted Games–Howell tests. Analyses used PyTorch [60], SciPy [61], Optuna [62] for the loss-weight sweep, and R for inferential statistics.

## 3 Results

Three results carry this section. First, the uncertainty map has a price, and it is small: the heteroscedastic likelihood buys the per-pixel aleatoric map for about 1.2 dB of PSNR. Second, for the VAE variant the KL-annealing schedule is decisive: cyclic annealing recovers a 1.10 dB gap over posterior-collapsing linear warmup, reaching statistical parity with the retrained CNN baselines. Third, the 1.2 dB price is not fundamental: two-stage fine-tuning closes it to a statistically indistinguishable 0.010 dB.

### 3.1 Reconstruction performance

Table 2 reports mid-noise reconstruction for the key arms and baselines. Because each test image carries fixed noise and each model returns a single reconstruction, the metrics are exactly repeatable, so sub-decibel gaps are genuine differences.

**Table 2:** Reconstruction and held-out perceptual metrics at mid noise (*n*=624, mean; PSNR ±SD). Best value per column in bold. Unc.: uncertainty maps produced (Ale. aleatoric, Epi. epistemic).

| Model | Unc. | PSNR (dB) | SSIM | FSIM↑ | LPIPS↓ |
| --- | --- | --- | --- | --- | --- |
| *Pixel-norm and fine-tuned arms* |  |  |  |  |  |
| Arm A (L2) | — | **$34.618 \pm 1.071$** | **0.9269** | 0.9317 | 0.2257 |
| Arm R (FT: L1+SSIM+FFL) | — | $34.545 \pm 1.067$ | 0.9269 | 0.9332 | **0.2192** |
| Arm S (FT: NLL+SSIM+FFL) | Ale. | $34.608 \pm 1.076$ | 0.9266 | 0.9317 | 0.2260 |
| Arm Q (Charbonnier) | — | 34.535 | — | 0.9315 | 0.2242 |
| Arm I (L1) | — | 34.444 | — | 0.9296 | 0.2291 |
| *Uncertainty arms (single-stage)* |  |  |  |  |  |
| Arm B (NLL) | Ale. | 33.46 | 0.909 | 0.9157 | 0.2732 |
| Arm D (NLL+SSIM+FFL) | Ale. | $33.435 \pm 1.026$ | 0.9089 | 0.9160 | 0.2733 |
| Arm H (PP-VAE, cyc+fb) | Ale.+Epi. | $32.762 \pm 1.064$ | 0.8957 | 0.9056 | 0.3079 |
| Arm F (PP-VAE, cyclic) | Ale.+Epi. | 32.73 | — | 0.9072 | 0.3009 |
| Arm E (PP-VAE, linear; collapsed) | Ale. | $31.635 \pm 0.951$ | 0.8726 | 0.8926 | 0.3687 |
| *Failure mode* |  |  |  |  |  |
| Arm O (PReLU) | — | $29.074 \pm 0.909$ | 0.8037 | 0.8757 | 0.4397 |
| *Retrained baselines (no uncertainty map)* |  |  |  |  |  |
| SCUNet | — | $34.149 \pm 1.094$ | 0.9127 | **0.9335** | 0.2201 |
| NAFNet | — | $33.803 \pm 1.100$ | 0.9068 | 0.9288 | 0.2305 |
| SharpXR | — | $33.561 \pm 1.107$ | 0.9019 | 0.9249 | 0.2368 |
| DRUNet | — | 32.58 | 0.866 | 0.9212 | 0.2354 |
| DnCNN | — | $32.798 \pm 0.941$ | 0.8774 | 0.9071 | 0.2922 |
| SwinIR | — | 31.18 | 0.853 | 0.8905 | 0.4148 |
| BM3D (oracle $\sigma$) | — | $30.716 \pm 0.929$ | 0.8056 | 0.8916 | 0.3716 |

The pixel-norm and fine-tuned arms hold the reconstruction ceiling (Arm A 34.62 dB, Arm S 34.61 dB, Arm R 34.55 dB), edging every retrained baseline: the strongest, SCUNet, reaches 34.15 dB despite its 57 M parameters, with NAFNet (116 M) and SharpXR further back. Adding the heteroscedastic NLL head costs about 1.2 dB (Arm B vs. A): the central price the rest of this section tests. The VAE bottleneck under cyclic KL (Arm H) trades a further 0.7 dB for epistemic uncertainty; the collapsed linear-KL Arm E and the PReLU failure Arm O mark the floor, and every trained arm except Arm O beats BM3D even with oracle noise knowledge, confirming that additive-white-Gaussian estimators are structurally inadequate for Poisson–Gaussian noise.

### 3.2 Held-out perceptual metrics

Neither FSIM nor LPIPS appears in any training objective, so both are fully held-out judges (Table 2, right columns). The broad ranking is preserved (rank correlation with PSNR *r*_s_ = 0.95), with one decisive exception: the perceptual optimum is not the PSNR optimum. Arm R posts the lowest LPIPS (0.2192) of any condition and the highest FSIM of any ablation arm (0.9332), while ranking only third on PSNR (−0.073 dB vs. Arm A).

One condition sits above it on FSIM: SCUNet (0.9335), a significant but negligible-sized gap (*d* = 0.19), which SCUNet concedes on LPIPS (*d* = 0.22); the two split the perceptual metrics. Arm R’s single-stage parent Arm J is inferior on both, so the advantage comes specifically from the two-stage schedule: an L1-trained checkpoint fine-tuned under a composite perceptual objective. This is the perception–distortion trade-off [63] resolved in the perceptual direction.

All conditions degrade monotonically across the three noise presets (Fig. 2), and the ranking is stable across the dose range (Spearman *r*_s_ *>* 0.98), so mid-noise comparisons are representative rather than an artefact of one operating point. Fig. 3 shows all 27 conditions across all four metrics at once.

**Figure 2:**
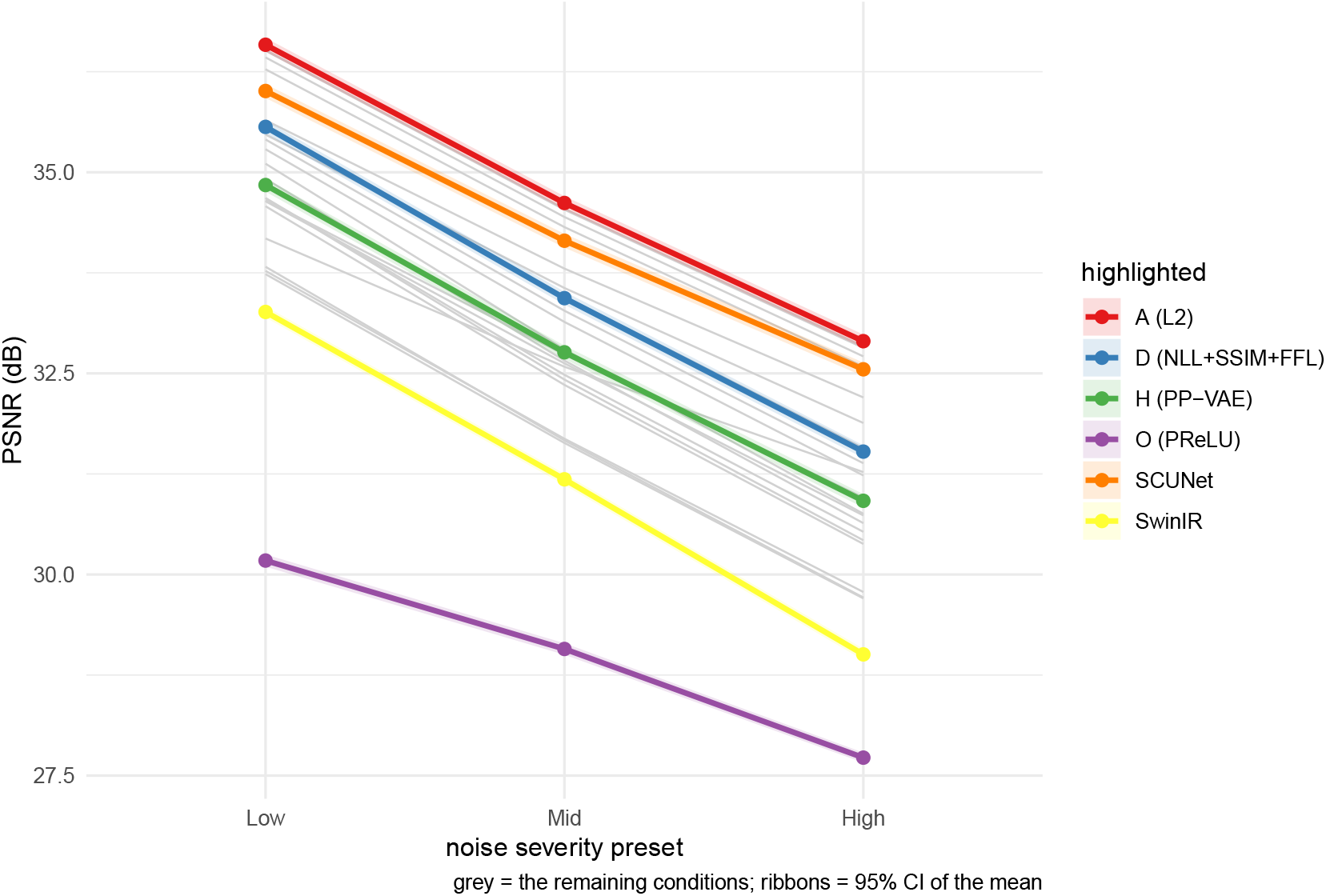
Mean PSNR versus noise-severity preset for all 27 conditions (19 arms and eight retrained baselines). Arm A leads at every dose ahead of SCUNet, the strongest retrained baseline; Arms D and H follow ≈1–2 dB below with their uncertainty maps; SwinIR trails the CNN baselines; Arm O (PReLU) collapses at every level. Six highlighted conditions carry 95% CI ribbons (*n*=624).

**Figure 3:**
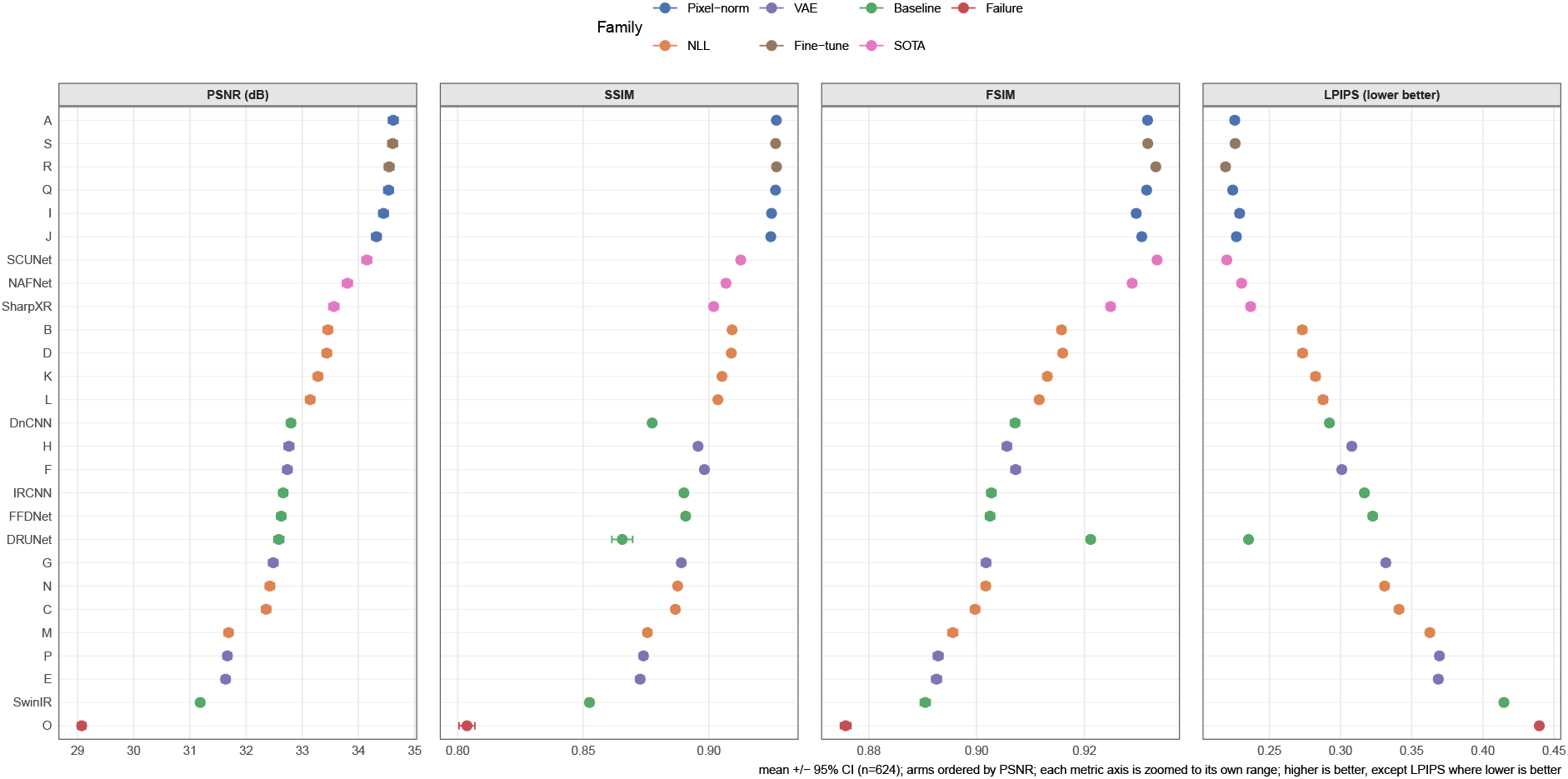
All 27 conditions across every metric at mid noise, ordered by PSNR and coloured by loss/architecture family (PSNR, SSIM, FSIM higher is better; LPIPS lower). The held-out FSIM and LPIPS preserve the family-level block structure of the PSNR ranking while reordering arms within blocks, so the broad ranking is not an artefact of any single objective. Error bars are 95% CIs (*n*=624).

### 3.3 Statistical comparisons

After an omnibus ANOVA rejects the global null, the Bonferroni-corrected pairwise Welch tests over all 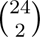 = 276 comparisons are summarised as Cohen’s *d* matrices in Fig. 4, one panel per metric; the marked non-significant cells identify the statistically interchangeable configurations, and the same loss-family block structure recurs under the held-out FSIM and LPIPS judges. The tests confirm six conclusions. (i) L1, L2 and Charbonnier are statistically indistinguishable in PSNR, SSIM and LPIPS (*d <* 0.2), consistent with all three pixel norms targeting the same posterior mean and over-smoothing similarly [17]. (ii) NLL alone and NLL+SSIM+FFL are indistinguishable in PSNR, but adding SSIM+Edge (Arm M) causes catastrophic gradient interference. (iii) Cyclic KL outperforms linear warmup by 1.10 dB; free bits added to cyclic is equivalent to cyclic alone. (iv) The VAE bottleneck itself imposes no detectable PSNR penalty: Arms E and F are architecturally identical, so the 1.10 dB gap reflects the schedule alone. (v) PReLU collapses catastrophically (*d* = +5.58 vs. Arm D). (vi) In PSNR the KAIR CNN baselines are indistinguishable from the cyclic-KL VAE arms, so the VAE adds calibrated uncertainty at no measurable PSNR cost relative to that family; the retrained SOTA baselines (NAFNet, SCUNet, SharpXR) form a higher cluster, still significantly below the pixel-norm and fine-tuned arms.

**Figure 4:**
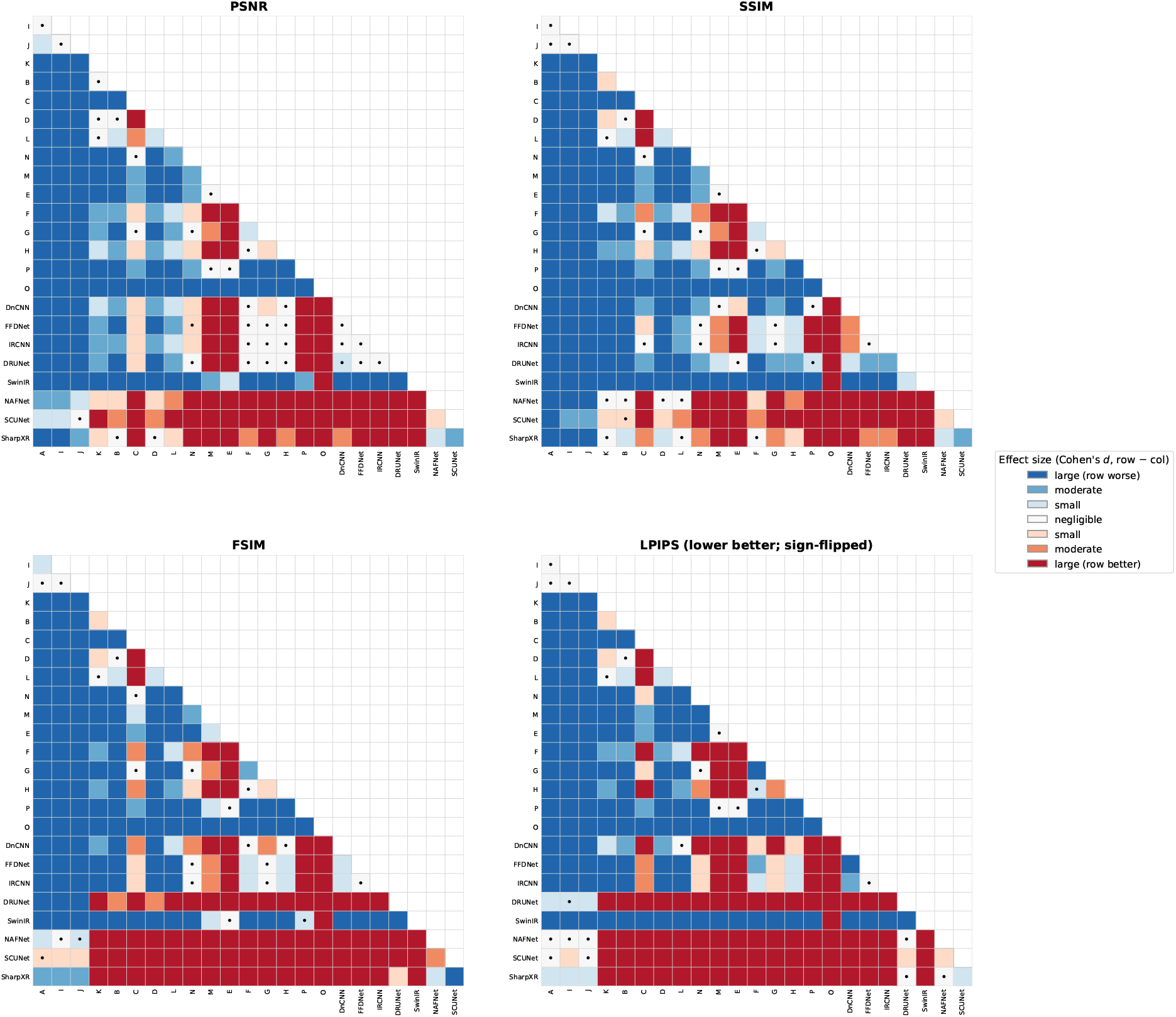
Pairwise Cohen’s *d* (lower triangle, row arm − column arm) for all 24 conditions (276 comparisons) at mid noise, one panel per metric: the trained metrics PSNR and SSIM (top) and the held-out perceptual judges FSIM and LPIPS (bottom). Red = row arm better, blue = worse (LPIPS sign-flipped so its colours read the same way). The palette is binned to the Cohen’s-*d* categories of Section 2.8: white = negligible (|*d*| *<* 0.2), pale = small (0.2–0.5), mid = moderate (0.5–0.8), saturated = large (≥ 0.8). A black dot marks a pair *not* significant after Bonferroni correction (statistically interchangeable). The same block structure recurs in every panel, so the ordering is not an artefact of any single objective.

### 3.4 Recovering the cost: two-stage fine-tuning

Table 3 and Fig. 5 summarise the two-stage loss-substitution arms. Neither fine-tuned arm is separable from the Arm A base (S vs. A: *t* = 0.17, *p* = 0.87, *d* = 0.01), while two-stage beats single-stage-from-scratch decisively (S vs. D: *t* = 19.7, *p <* 10*^−^*^70^, *d* = 1.11). Arm S therefore delivers the calibrated aleatoric map (NLL −2.650, sharpness 0.0201) at the L2 ceiling: the 1.2 dB calibration tax is a training-trajectory artefact, not a fixed cost.

**Figure 5:**
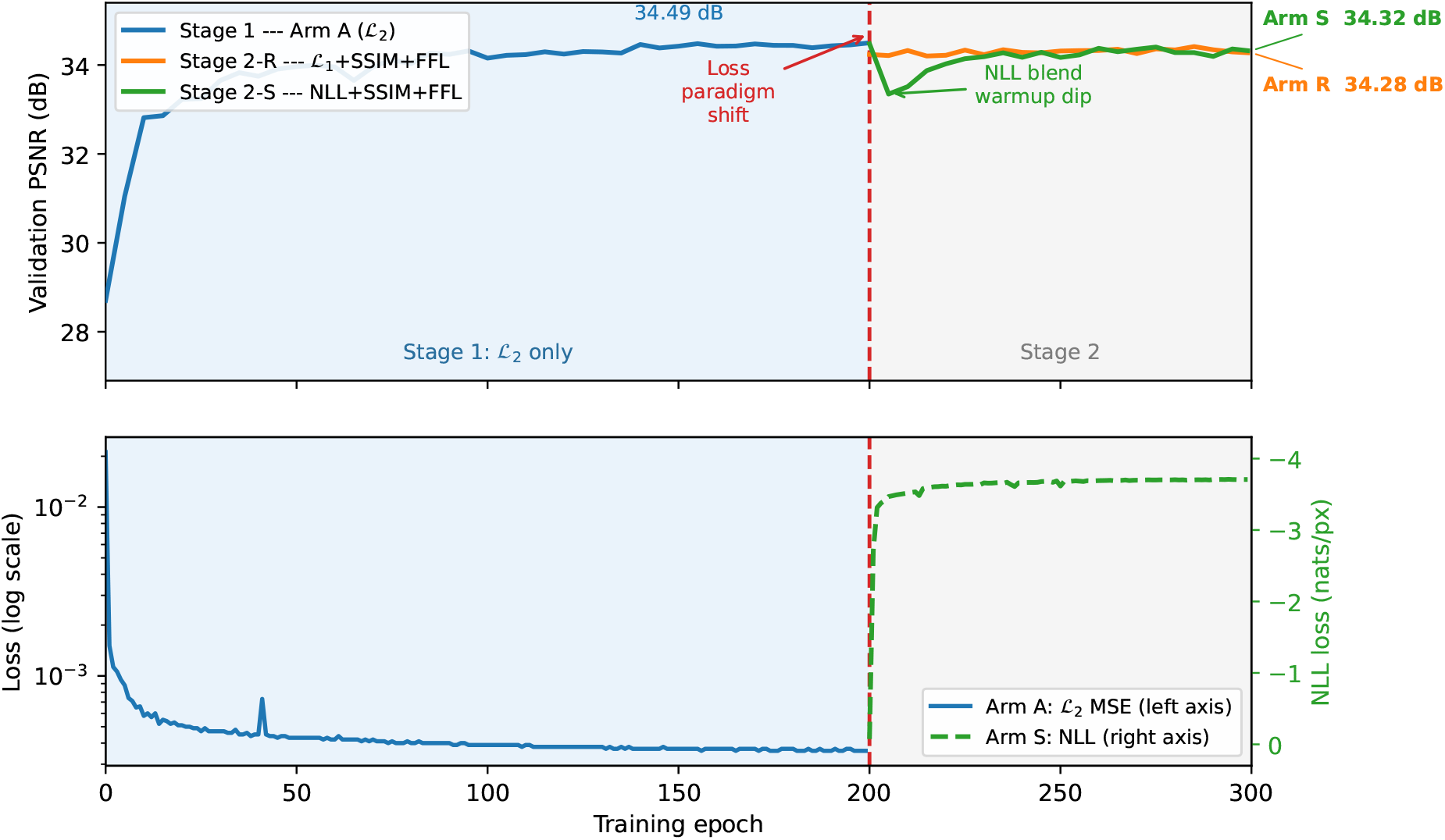
Two-stage fine-tuning dynamics. Top: 300-epoch validation-PSNR trajectory: Arm A converges on MSE (epochs 0–199), then Arms R and S resume from its weigh ts under a composite loss (dashed line at epoch 200); Arm R holds Arm A’s level throughout, while Arm S dips as its variance head re-stabilises and recovers within ≈40 epochs. Bottom: corresponding training losses (MSE left axis, NLL right axis).

**Table 3:** Two-stage loss-substitution fine-tuning vs. single-stage counterparts at mid noise.

| Arm | Objective | PSNR (dB) | LPIPS↓ | ΔPSNR vs. A |
| --- | --- | --- | --- | --- |
| A | L2 (Stage-1 base) | $34.618 \pm 1.071$ | 0.2257 | — |
| R | L1+SSIM+FFL (Stage 2) | $34.545 \pm 1.067$ | **0.2192** | −0.073 dB |
| S | NLL+SSIM+FFL (Stage 2) | $34.608 \pm 1.076$ | 0.2260 | −0.010 dB |
| J | L1+SSIM+FFL (scratch) | $34.319 \pm 1.060$ | 0.2268 | −0.299 dB |
| D | NLL+SSIM+FFL (scratch) | $33.435 \pm 1.026$ | 0.2733 | −1.183 dB |

### 3.5 Qualitative and region-of-interest assessment

Fig. 6 shows Arm H on one representative case per diagnostic class. The denoised mean restores the anatomy the noise hid; the aleatoric map is bright along rib, vessel and consolidation borders and dark over smooth lung; and the *σ̂*_a_ and error columns light up in the same places: the visual form of the pixel-level calibration quantified below. The epistemic column flags a partly different set of regions (consolidation and cardiac borders), and exists only because the cyclic schedule keeps the latent active.

**Figure 6:**
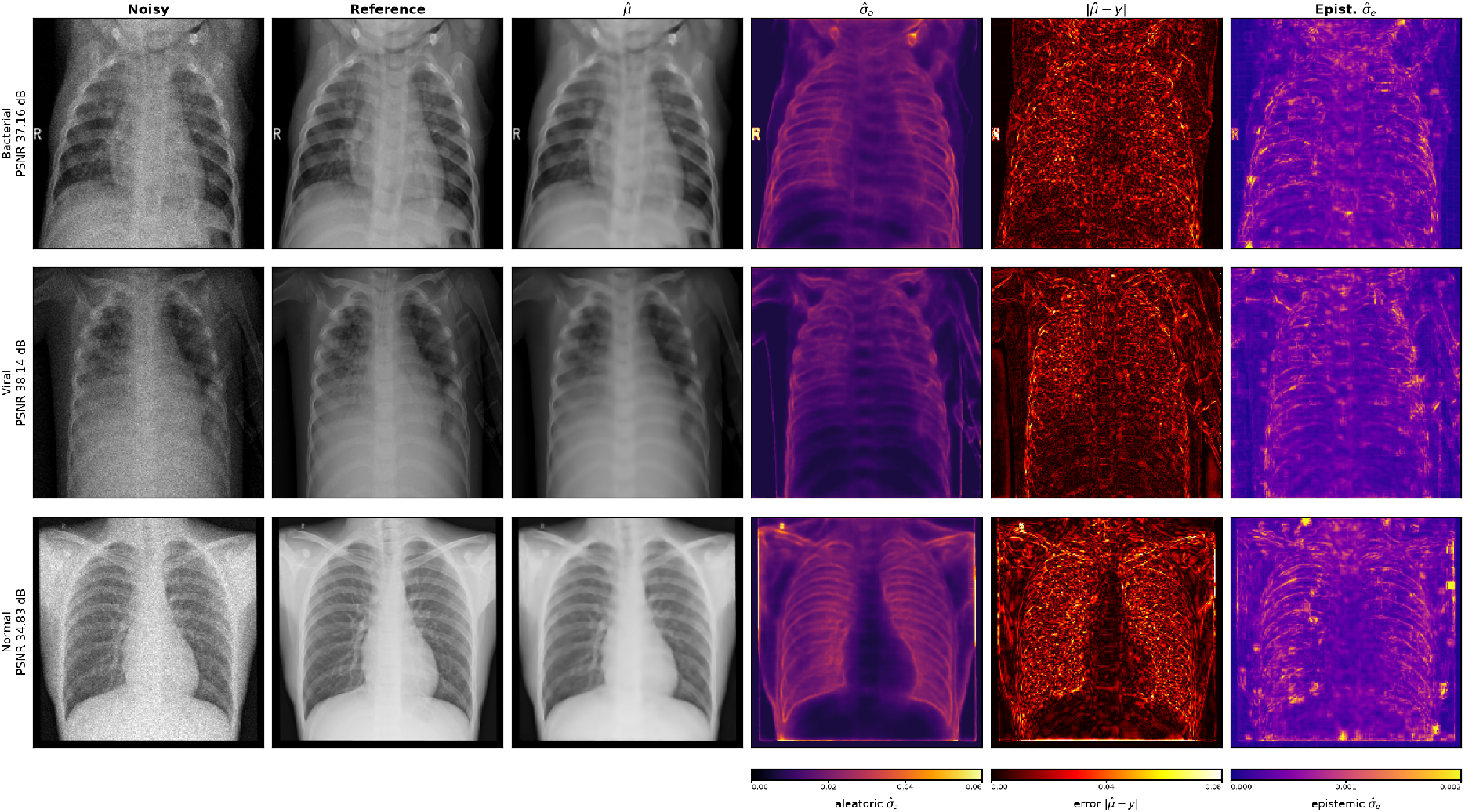
Arm H (best VAE) on one representative case per class at mid noise. Columns: noisy input, clean reference, denoised mean *µ̂*, aleatoric map *σ̂*_a_ (inferno, shared 0–0.06 scale), absolute error |*µ̂* − *y*|, epistemic map *σ̂*_e_ (plasma, own scale). Epistemic uncertainty localises to consolidation boundaries (Bacterial) and cardiomegaly margins (Viral), staying low in Normal cases.

Region-of-interest crops at the four hardest, most diagnostically decisive sites (fine pulmonary vessels, posterior rib cortical margin, costophrenic angle, perihilar vessels; Fig. 7) show that *no* condition (ours or the far larger SCUNet and NAFNet) resolves any site back to the reference; the objective sets only how soft the residual looks. The frequency-aware arms retain high-frequency hilar and vascular texture that mean-seeking losses smooth away, which is why Arm R attains the best LPIPS at a sub-decibel PSNR cost. SwinIR produces a faint periodic grid at all four sites, most concerning at the costophrenic angle where it could mimic early effusion blunting.

**Figure 7:**
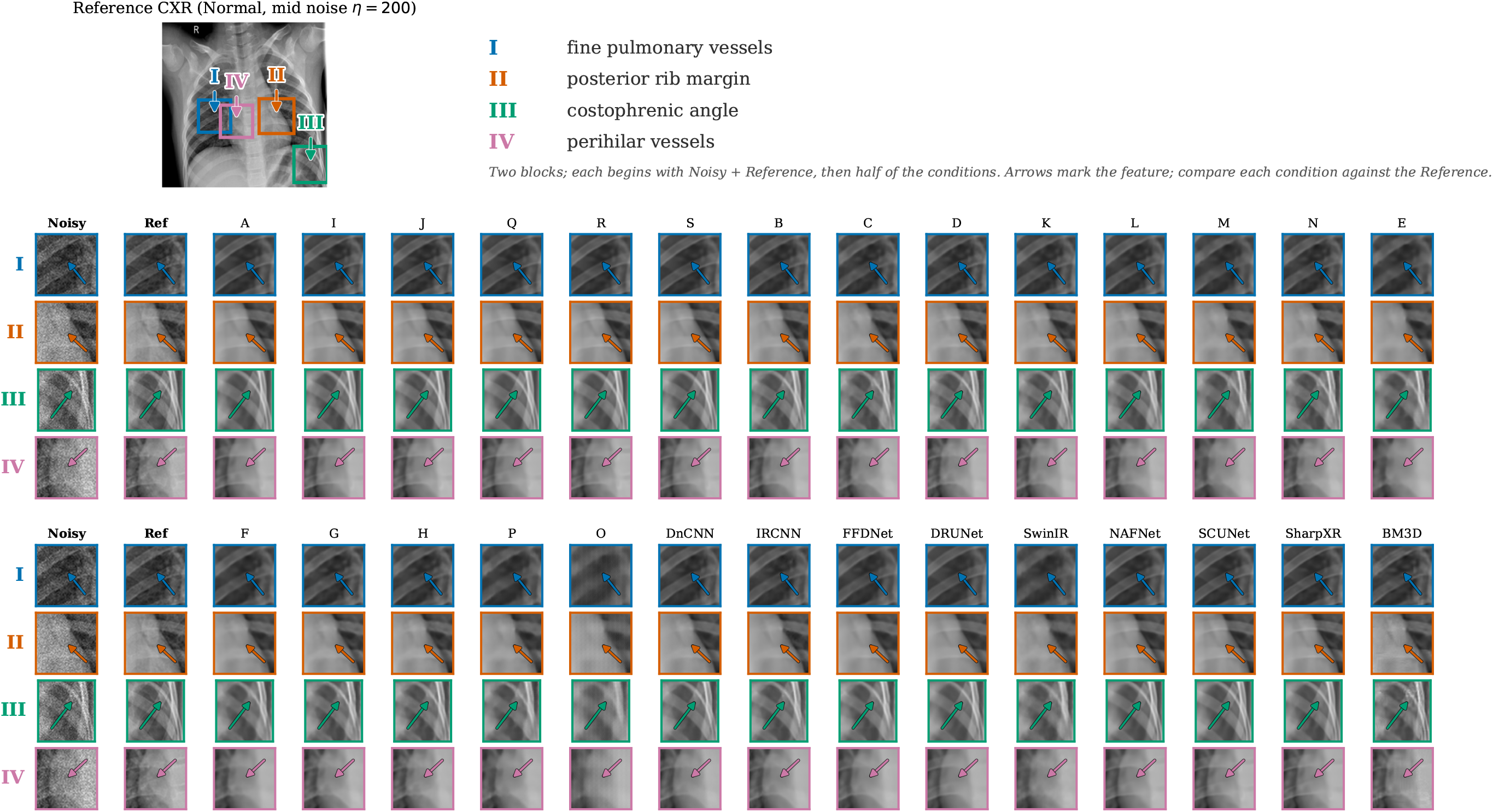
Region-of-interest comparison at mid noise. Top left: reference CXR with four arrowed sites (I fine vessels, II posterior rib margin, III costophrenic angle, IV perihilar vessels). Below: zoomed crops for every ablation arm and all eight retrained baselines. No model recovers the finest vessels to full realism; differences are of degree, not kind.

### 3.6 Sensitivity to loss weights

A 14-trial Optuna sweep of Arm D’s composite weights finds a smooth, single-peaked validation-PSNR optimum (*λ*_SSIM_ ≈ 0.96, *λ*_FFL_ ≈ 0.02) about 1.2 dB above the literature-default weights used throughout, so the reported ablation metrics are conservative; all four metrics agree on the optimum (Spearman *r*_s_ ≥ 0.89). A supplementary edge/SSIM sweep finds no re-weighting that recovers Arm M.

### 3.7 Latent space

The bottleneck is the source of the epistemic map, so whether its KL schedule trained the latent rather than collapsing it underwrites that signal. *k*-means clustering of the pooled posterior means (*n* = 448 test images, *k* = 3) shows the schedule is the primary driver of class structure: Arm F (cyclical) reaches adjusted Rand index 0.333 [0.242, 0.429] and silhouette 0.113 with visible class regions in t-SNE/UMAP projections (Fig. 8), Arm H (cyc+fb) 0.210, while the collapsed Arm E gives ARI −0.002 and free-bits-only 0.048. Arm F separates classes better while Arm H reconstructs marginally better, a structure-versus-reconstruction choice rather than a single best setting.

**Figure 8:**
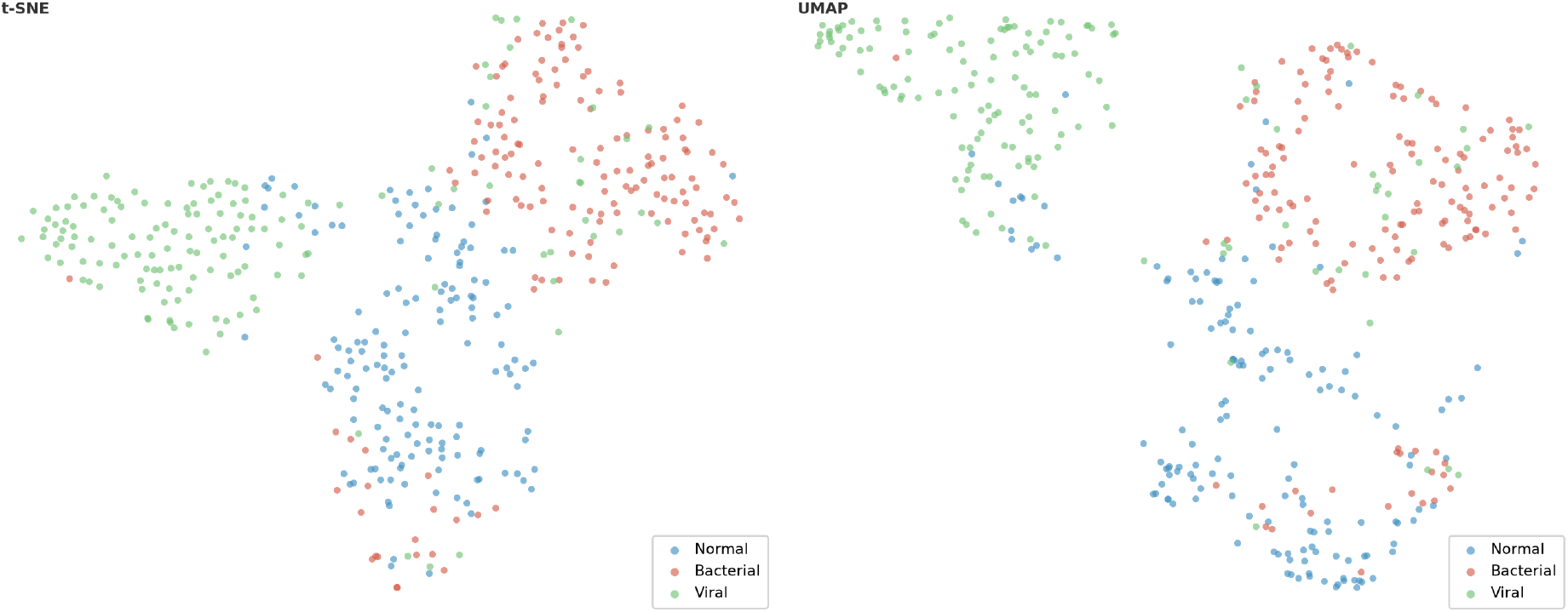
t-SNE (left) and UMAP (right) of Arm F latent codes (*n*=448), coloured by ground-truth class. Class-structured regions emerge although the model sees no class labels (ARI = 0.333, the highest of the five VAE arms); Viral forms the most compact cluster, Normal and Bacterial partially separate.

### 3.8 Calibration

Arms B and D achieve NLL ≈ −2.64 nats/pixel at mid noise (Arm H ≈ −2.55), sharpness 0.020 (B/D) and 0.022 (H). Reliability diagrams (Fig. 9) show observed RMS error tracking predicted *σ̂*_a_ along the diagonal (Pearson *r* ≈ 0.8–0.9), and the post-hoc *σ*-scaling puts the size on a formal footing: the optimal scale is within 1% of unity (*s*^⋆^ = 1.005, 1.008, 1.002 for Arms B, D, H) with mean absolute calibration error ≈ 0.001, essentially unchanged by rescaling. A predicted standard deviation can therefore be read close to face value: a pixel flagged *σ̂* ≈ 0.02 carries an error of about 0.02, not merely more than one flagged 0.01. The map also behaves as a dose-response signal: mean *σ̂̄*_a_ rises monotonically as dose falls (Arm D 0.020 → 0.031; Arm H 0.022 → 0.034), with Arm H’s epistemic map climbing in parallel (0.0006 → 0.0011), consistent with the model leaning more on its prior as the measurement weakens.

**Figure 9:**
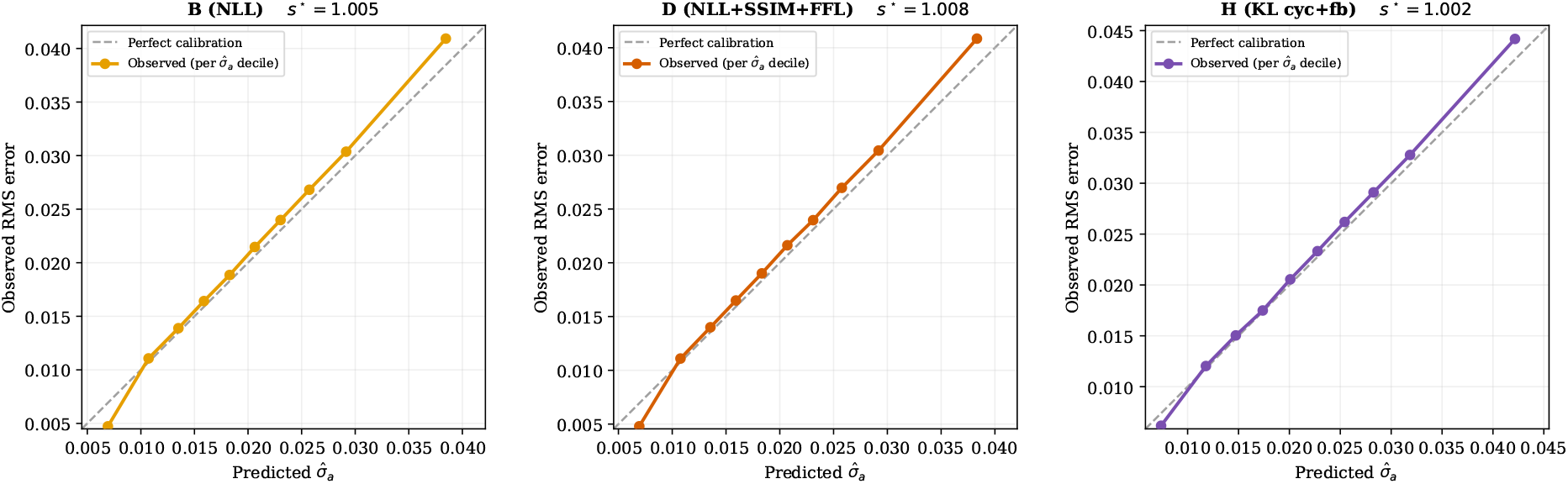
Pixel-level reliability diagrams for Arms B, D and H at mid noise: held-out test pixels grouped into deciles of predicted *σ̂*_a_, each decile’s observed RMS error plotted against its mean prediction; the dashed diagonal is perfect calibration. All three track the diagonal closely, with the single post-hoc scale *s*^⋆^ within 1% of unity (annotated per panel). The inverse-crime caveat of the Discussion applies.

#### Aleatoric and epistemic maps are complementary

For Arm H the aleatoric map is ∼30× larger in magnitude and concentrates on the high-contrast bony thorax, consistent with re-expressing the injected Foi noise; the epistemic map is only moderately correlated with it (per-image Pearson *r̄* = 0.53 ± 0.07 over 100 images; top-decile Jaccard 0.21 ± 0.03; Fig. 10). The epistemic map is not a rescaled copy: the aleatoric channel answers “how noisy is this pixel” and the epistemic channel “how much of this pixel is invented by the model.”

**Figure 10:**
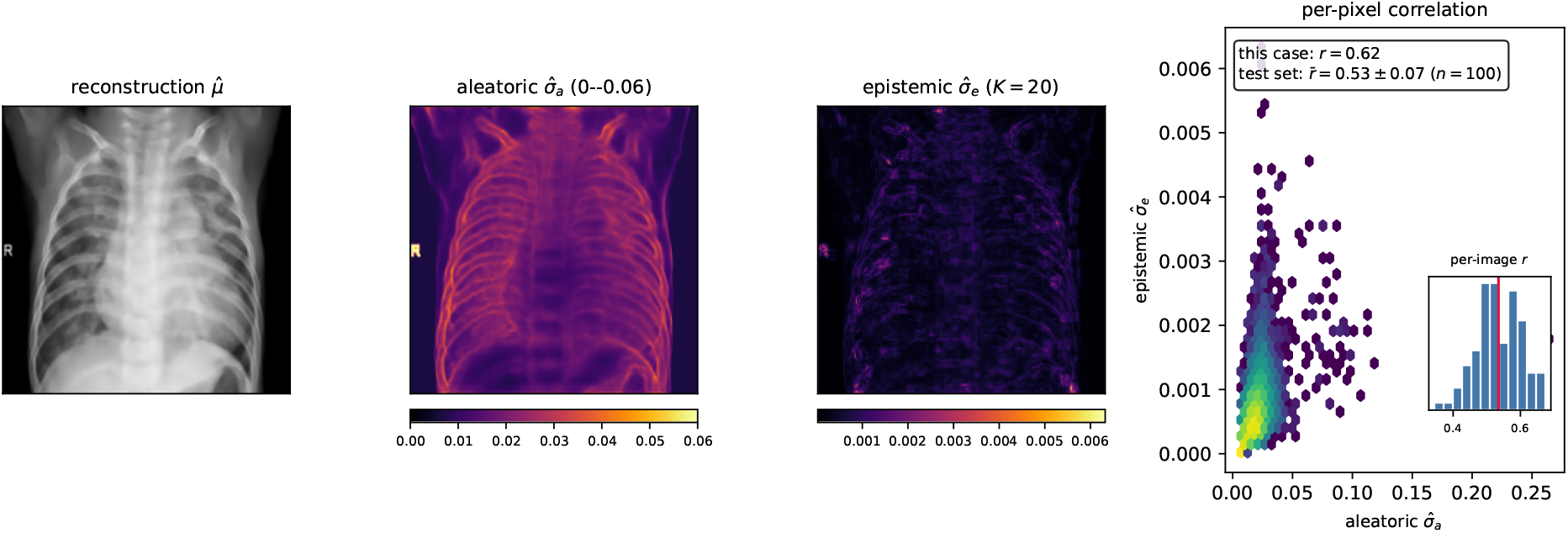
Aleatoric and epistemic uncertainty are complementary (Arm H, one Bacterial case). Left to right: reconstruction *µ̂*; aleatoric *σ̂*_a_ (shared 0–0.06 scale, concentrated on the bony thorax and edges); epistemic *σ̂*_e_ (*K*=20 samples, own scale, ∼30× smaller and spatially distinct); per-pixel correlation panel (this case *r* = 0.62; inset: distribution of per-image *r* over 100 images, *r̄* = 0.53 ± 0.07).

### 3.9 Breaking the inverse crime: a cross-degradation stress test

The calibration above is measured on the same Foi noise the models were trained on, so it cannot by itself distinguish genuine calibration from the inverse crime [19]. We therefore re-evaluate the fixed checkpoints (inference only, no retraining) on the identical 624-image test set corrupted by six degradations, each *calibrated to the same input PSNR as the training noise* (17.5 dB) so that differences reflect the noise *model*, not its magnitude: two draws from the training family (a shifted read-noise floor and shifted (*a, b*)), three genuinely different processes at the same severity (pure Poisson shot noise, mild blur plus noise, and multiplicative speckle), and JPEG compression, which is excluded from the analysis below because its PSNR floors near 22 dB even at quality 1 and so cannot be matched to the noisier anchor (Fig. 11).

**Figure 11:**
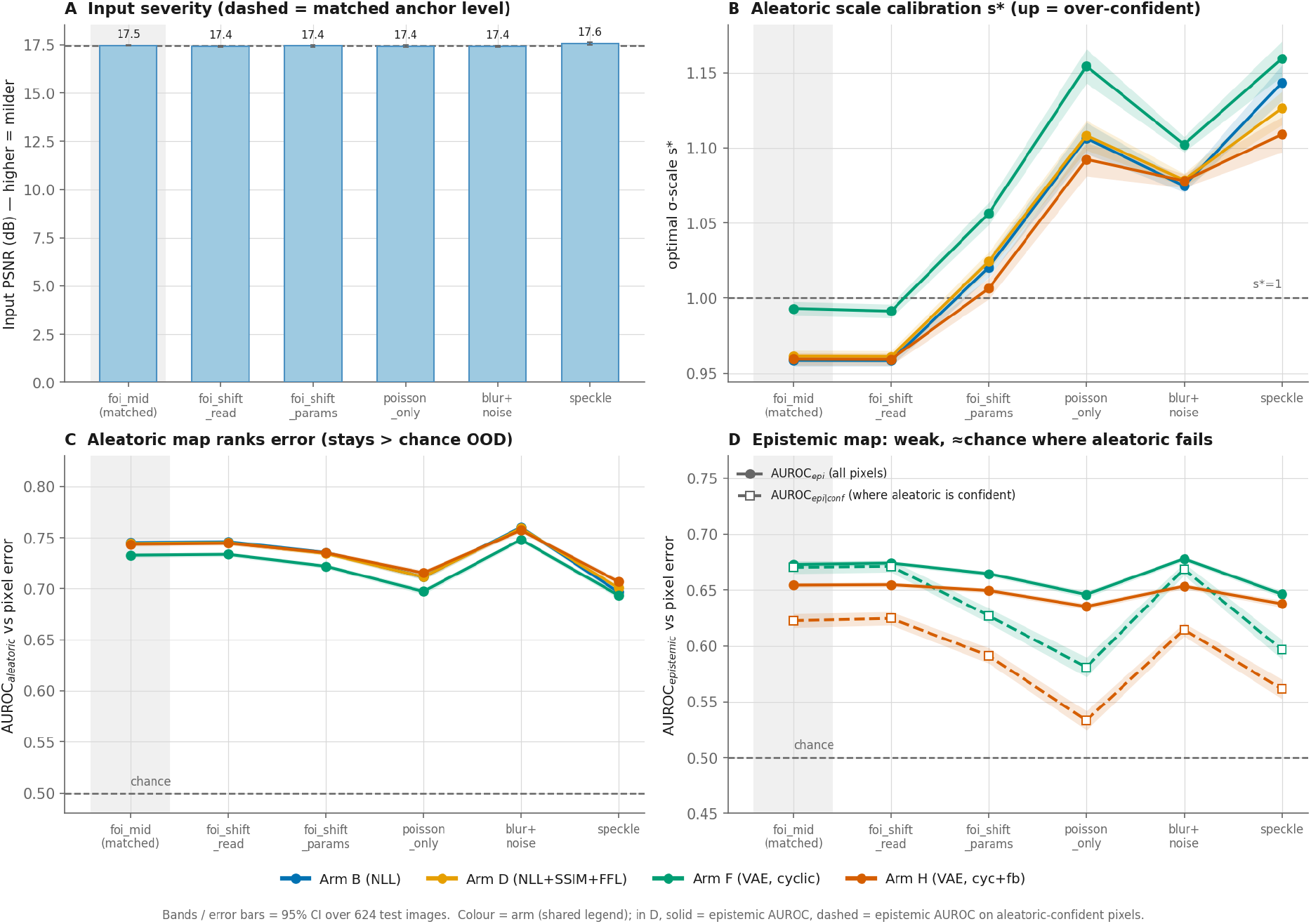
Cross-degradation stress test at matched severity (every degradation calibrated to 17.5 dB input PSNR; *n* = 624; inference-only). **A** input severity, confirming the degradations are equalised. **B** aleatoric scale calibration *s*^⋆^ (1 = calibrated, *>* 1 = over-confident): calibrated under same-family shifts (foi shift read/params) but rising to 1.08–1.16 once the noise *model* changes (Poisson-only, blur+noise, speckle), the direct signature of the inverse crime. **C** the aleatoric map’s error-*ranking* (AUROC) stays well above chance throughout. **D** the epistemic map is a weaker error predictor than the aleatoric map everywhere, and its confident-but-wrong AUROC (dashed) drops toward chance under the model mismatches. Bands are 95% CIs over the 624 images.

Two results follow. First, the aleatoric map’s calibration is model-circular. It stays calibrated under the same-family shifts (*s*^⋆^ = 0.96–1.02, matching the in-distribution anchor) but becomes *modestly but systematically* over-confident once the noise model changes at equal severity: *s*^⋆^ rises to 1.08 (blur+noise), 1.11 (Poisson-only) and 1.13 (speckle) for the aleatoric arms (and to 1.16 for the variational arms), meaning realised errors are 8–16% larger than the map predicts, with the calibration correlation falling from 0.995 to ≈ 0.97. The shift is small in absolute terms but tightly bounded (the 95% CIs exclude the same-family control), so it is a genuine, if modest, signature rather than noise; and because the mismatches tested are mild, severity-matched, and summarised by a single global scale, it is best read as a *lower bound* on the over-confidence a real acquisition shift (or a local, low-contrast region) could induce. Its *ranking* of error is by contrast robust (AUROC of *σ̂*_a_ against thresholded pixel error stays 0.69–0.76 across every degradation): the map still flags where errors concentrate, but mis-states their size out of distribution. Second, the epistemic map does not compensate. Its error-ranking AUROC (0.63–0.68) is below the aleatoric map’s in every condition, and its ability to catch *confident-but-wrong* pixels (those the aleatoric map calls confident yet that carry high error) is weakest precisely under the model mismatches where the aleatoric map most over-confidently fails, falling from 0.62 in-distribution to 0.53 (Poisson-only) and 0.56 (speckle) for Arm H, approaching chance. Under matched go for severity, therefore, Monte-Carlo epistemic uncertainty is not a reliable substitute for the aleatoric map where the latter breaks.

### 3.10 Fairness across diagnoses

No objective is biased against the pathological classes: both pneumonia classes reconstruct at higher PSNR than Normal in every model, by a nearly constant margin (Bacterial +1.1, Viral +0.9 dB), because lobar consolidation replaces fine vascular detail with smooth opacity that is easier to denoise; the ordering is a property of the images, not the models, and the held-out LPIPS agrees.

### 3.11 Failure modes as design lessons

Three arms fail instructively (Table 4), and share a lesson about heteroscedastic training: PReLU’s learnable negative slope lets the log-variance head diverge (variance-predicting layers need bounded activations such as GELU); linear KL warmup collapses the posterior to a deterministic decoder with structureless epistemic maps; and SSIM and Sobel-edge terms pressure the same spatial gradients through conflicting pathways: a previously unreported interaction invisible to either term alone and unrecovered by any re-weighting.

**Table 4:** Failure modes with mechanism and PSNR impact at mid noise (vs. Arm D).

| Arm | Failure | Mechanism | PSNR |
| --- | --- | --- | --- |
| O | PReLU instability | log-variance head diverges (epoch 12) | 29.07 dB (−4.36) |
| E | Posterior collapse | linear KL warmup; $D_{KL} < 0.01$ nats | 31.64 dB (−1.80) |
| M | Gradient interference | SSIM + Sobel supervise the same gradients | 31.69 dB (−1.75) |

## 4 Discussion

### The answer to the research question is yes

The calibrated per-pixel uncertainty map is obtained at effectively no cost to reconstruction quality. The heteroscedastic head’s 1.2 dB price is a training-trajectory artefact, not a ceiling: two-stage fine-tuning closes it to 0.010 dB, statistically indistinguishable from the L2 base. The same fine-tuning resolves the perception–distortion trade-off [63] in the perceptual direction, with Arm R and SCUNet splitting the two held-out perceptual judges without either leading PSNR.

### Two levers matter more than parameter count

Among the retrained CNN baselines, years of architectural change buy little (DnCNN through DRUNet sit within 0.22 dB of one another). The first lever is the backbone: the hybrid convolution–attention design exceeds DnCNN by 1.82 dB at the identical L2 loss, at 25.7 M parameters against SCUNet’s 57 M and NAFNet’s 116 M, so efficient attention rather than scale sets the ceiling. The second lever is the objective on that fixed backbone: the KL schedule alone is worth 1.10 dB, the SSIM+FFL terms buy perceptual quality a pixel loss discards, and the NLL head buys the calibrated map at a cost fine-tuning erases. The NLL+SSIM+FFL objective is architecture-agnostic: it requires only a (mean, log-variance) decoder head and transfers to any heteroscedastic encoder–decoder. A cautionary corollary: pure windowed self-attention (SwinIR), competitive on additive-Gaussian benchmarks, trails plain CNNs under Poisson–Gaussian paediatric CXR noise.

### An information limit, not a capacity limit, bounds every denoiser

At the diagnostically decisive regions no model recovers the finest vessels to full realism: once low-dose noise buries a thread-like vessel, by the data-processing inequality [64, §2.8] no post-processing (deterministic or stochastic) can increase the information the measurement retains about the underlying anatomy. What a denoiser does next is the forced choice of the perception–distortion trade-off: a mean-seeking objective smooths the missing detail into blur [17], whereas a generative denoiser [11, 12, 65] paints in a sharp, plausible hallucination [13, 14], arguably the more dangerous failure for a diagnostic image. A calibrated uncertainty map is precisely the instrument that makes this limit visible per pixel.

### What the uncertainty map is, and is not

The map’s meaning must be read with care. Because the degradation is simulated with the Foi model and the heteroscedastic head is trained to predict *ay* + *b*, the aleatoric map largely re-expresses the injected noise (the inverse crime [19]), so its near-perfect calibration is partly circular, and in contrast-to-noise terms it flags high-contrast structure rather than the low-contrast findings the Rose criterion marks as most vulnerable [4]. Our matched-severity cross-degradation test (Section 3.9) makes this circularity concrete: holding input PSNR fixed, the aleatoric map stays calibrated under same-family noise but becomes modestly (8–16%) over-confident once the noise *model* changes, while still *ranking* error reliably. The effect is small but systematic, and a lower bound: the shifts tested are mild and severity-matched, so a real acquisition change could induce more. The same test tempers the epistemic map: although it is untethered from the injected noise and only moderately correlated with the aleatoric one, it is a weaker error predictor in every condition and does *not* compensate where the aleatoric map fails, so it is best read as a complementary flag for prior-driven reconstruction rather than a calibrated substitute. Closing this gap, an architecture-independent epistemic estimate such as a deep ensemble [34], and validation on real low-dose acquisitions, is the priority next step.

### Deployment profiles

Two configurations follow from the results. Arm S is the aleatoric-triage configuration: a single sub-50 ms forward pass returns the calibrated *σ̂*_a_ map at the full L2-level fidelity. Arm H adds the epistemic channel when needed, at the cost of *K*=20 stochastic passes (≈1 s); its epistemic map is a candidate distribution-shift signal of the kind recent paediatric-AI reviews call for [15], though our stress test (Section 3.9) shows it is only weakly predictive and would need strengthening (e.g. deep ensembling) before clinical use. Translating the high-noise operating points (Arm A holds 32.9 dB at roughly 3× reduced dose) into a dose-reduction claim requires a prospective reader study.

### 4.1 Limitations and future work

The clearest limitation is that the degradation is simulated in the image domain, so noise scales with displayed brightness and falls hardest on bone rather than on the photon-starved lung; a projection-domain model on raw data would be the single most improving change, and even then a faithful aleatoric map would relocate to the photon-starved anatomy, leaving low-contrast findings to the epistemic channel. The single-centre Kermany data is JPEG-compressed and demographically narrow, so generalisation to other collections (MIMIC-CXR [66], CheXpert) is unestablished; the 62.5/37.5 pneumonia/normal split inflates aggregate PSNR by ≈0.4 dB without changing rankings; labels are binary, precluding severity analysis; loss weights were not exhaustively searched; the three noise presets are fixed reference points rather than a sample of the clinical dose distribution; and the VAE findings rest on a single backbone, so the schedule-over-architecture conclusion is untested across other encoder–decoders. No pixel metric substitutes for a radiologist, and no reader study was performed.

Future work, in order of clinical impact: a blinded radiologist reader study on Arms A, D and H (diagnostic confidence, lesion detection, and whether the uncertainty overlay supports reader decisions); validation on real, full-resolution low-dose scans (e.g. MIMIC-CXR-JPG [67]); anatomy-attributed, thresholded per-scan uncertainty reports (Fig. 12 sketches this overlay for Arm H, with threshold calibration against confirmed pathology the immediate next step); and, methodologically, *β*-TCVAE latent structure [68], a CXR-pretrained perceptual loss to replace the ineffective ImageNet VGG-16 [69], and deep ensembling [34] as an architecture-independent epistemic baseline.

**Figure 12:**
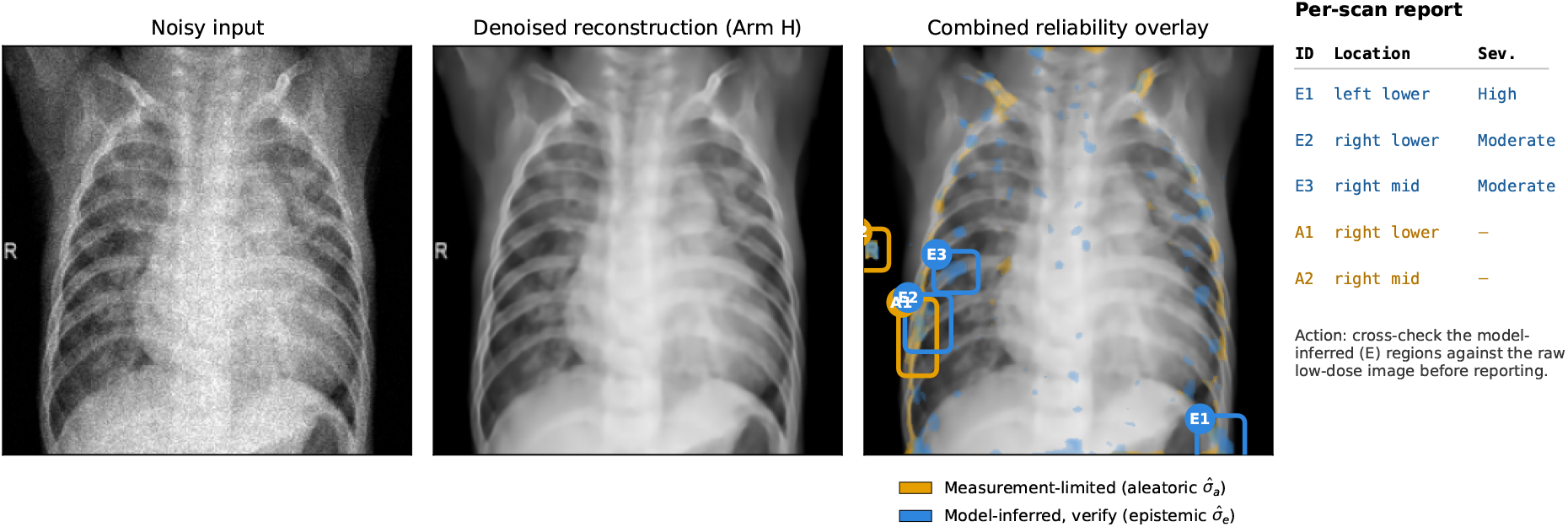
Prototype reliability-triage overlay (Arm H) on one representative case: noisy low-dose input, denoised reconstruction, and a combined overlay carrying both uncertainty channels with zone-attributed bounding regions and a per-scan report. Measurement-limited regions (amber, aleatoric) track the bony thorax; model-inferred regions (blue, epistemic, “verify”) mark where the latent prior filled the reconstruction in and are the clinically actionable signal. A design sketch, not a validated tool.

## 5 Conclusions

PP-VAE-Hformer inserts a variational bottleneck and a heteroscedastic dual head into a hybrid convolution–transformer denoiser and trains with a composite NLL+SSIM+FFL+KL objective. Across a 19-arm ablation and eight retrained baselines on Poisson–Gaussian-degraded paediatric chest radiographs, it shows that a calibrated per-pixel uncertainty map can be attached at effectively no cost to reconstruction quality: the heteroscedastic head’s 1.2 dB price is erased by two-stage fine-tuning (0.010 dB, statistically indistinguishable), the KL-annealing schedule rather than the bottleneck decides whether the variational variant reaches CNN parity, and the aleatoric map is calibrated to within 1% of unity under the training noise. A matched-severity cross-degradation test qualifies that last point: the calibration is tied to the trained noise model: the aleatoric map turns modestly (8–16%, a lower bound) over-confident once the noise model changes at equal severity, and the Monte-Carlo epistemic map, while flagging complementary regions, is a weaker error predictor that does not reliably substitute where the aleatoric map fails. For low-dose paediatric chest radiography, the confidence of a reconstruction can thus be made as reportable as the reconstruction itself *under a known noise model*; what stands between this and the clinic is validation: an architecture-independent uncertainty estimate such as deep ensembling, a blinded reader study, and real low-dose data, not the reconstruction method.

## Abbreviations

ARI: adjusted Rand index; CNN: convolutional neural network; CXR: chest radiograph; FFL: focal frequency loss; FSIM: feature similarity index; FT: fine-tuning; GELU: Gaussian error linear unit; KL: Kullback–Leibler (divergence); LPIPS: learned perceptual image patch similarity; MSE: mean squared error; MS-SSIM: multi-scale structural similarity; NLL: negative log-likelihood; PReLU: parametric rectified linear unit; PSNR: peak signal-to-noise ratio; ROI: region of interest; SSIM: structural similarity index; VAE: variational autoencoder.

## Declarations

### Ethics approval and consent to participate

Not applicable. This study used only the publicly available, fully de-identified Kermany paediatric chest radiograph collection [49]; no new human data were collected.

### Consent for publication

Not applicable.

### Availability of data and materials

The dataset supporting the conclusions of this article is the publicly available Kermany paediatric chest X-ray collection [49] (https://data.mendeley.com/datasets/rscbjbr9sj/2). The full training, evaluation and analysis code supporting the conclusions of this article is available in the PPVAE-Hformer-codebase repository, https://github.com/USN4730/PPVAE-Hformer-codebase.

### Authors’ contributions

STM and KF conceived the study. STM designed the study, implemented the models, performed the experiments and statistical analyses, and drafted the manuscript. KF supervised the work and revised the manuscript. Both authors read and approved the final manuscript.

## Data Availability

The dataset supporting the conclusions of this article is the publicly available Kermany paediatric chest X-ray collection

https://data.mendeley.com/datasets/rscbjbr9sj/2

## Acknowledgements

Model training used the Cambridge Service for Data Driven Discovery (CSD3).

